# Dynamic Imaging Markers of ARIA-H Risk in the A4 Study: A Person-Interval Analysis of Current Lesion Burden and Recent Microhaemorrhage Activity

**DOI:** 10.64898/2026.05.30.26354521

**Authors:** Callum Hill, Huw Morgan, Sofia Michopoulou, Mahesan Niranjan, Christopher M. Kipps

**Affiliations:** Clinical and Experimental Sciences, Faculty of Medicine, University of Southampton, Southampton, UK; School of Electronics and Computer Science, University of Southampton, Southampton, UK; Department of Medical Physics, University Hospital Southampton NHS Foundation Trust, Southampton; University Hospital Southampton NHS Foundation Trust, Southampton, UK

**Keywords:** ARIA-H, Alzheimer’s disease, microhaemorrhage, superficial siderosis, amyloid-related imaging abnormalities, A4 Study, anti-amyloid therapy, longitudinal MRI, risk prediction, solanezumab

## Abstract

**INTRODUCTION:** Amyloid-related imaging abnormalities with microhaemorrhage (ARIA-H) are an important safety consideration with anti-amyloid therapies, yet evidence exploring short-term risk prediction remains limited.

**METHODS:** Models incorporated baseline covariates, alongside dynamic variables of recent microhaemorrhage accumulation and current microhaemorrhage burden. Incident ARIA-H was defined as 2 or more new microhaemorrhages or 1 or more new superficial siderosis between consecutive MRI scans.

**RESULTS:** Among 1,069 participants (3,647 intervals), 171 ARIA-H events occurred. Both current burden (time-to-event: OR 1.37, 95% CI 1.09-1.73; all-event: OR 1.24, 95% CI: 1.04-1.49) and recent microhaemorrhage accumulation (time-to-event: OR 1.83, 95% CI 1.01-3.30; all-event: OR 1.43, 95% CI: 1.00-2.04) were independently associated with an increased risk of ARIA-H.

**DISCUSSION:** Temporal imaging variables may provide independent prognostic information beyond baseline risk, supporting a dynamic model of haemorrhagic risk in Alzheimer’s disease.

## 1 Introduction

Amyloid-related imaging abnormalities with microhaemorrhage (ARIA-H) are commonly observed in Alzheimer’s disease and in clinical trials of anti-amyloid therapies. [1,2] ARIA-H comprises microhaemorrhages (MCH) and superficial siderosis (SS), and is most commonly detected on MRI, particularly on GRE/T2-weighted sequences. [3] Although the exact aetiology remains unclear, current literature links ARIA-H to cerebral amyloid angiopathy (CAA), vascular fragility, and amyloid clearance across vessel walls. [2,4,5]

Given the increasing use of anti-amyloid therapies, ARIA-H has become a clinically relevant safety concern. [6,7] While often asymptomatic, ARIA-H represents a treatment safety signal as its occurrence reflects heightened haemorrhagic vulnerability. [5] The accumulation of ARIA-H, if of sufficient severity, can lead to treatment interruption or discontinuation, as well as increased MRI monitoring burden. [8,9] Identifying individuals at elevated risk of ARIA-H is of regulatory importance, given the need to balance treatment risks and benefits. [9,10] Despite its importance, literature examining the prediction of short-term risk of ARIA-H remains limited.

Existing studies have identified baseline risk factors associated with the development of ARIA-H, including baseline microhaemorrhage burden, APOE ε4 carrier status, vascular risk factors, and amyloid burden. [2, 11–13] Baseline microhaemorrhage count has been identified as a risk factor for ARIA-H development, likely reflecting existing cumulative vascular injury. [11] Genetic predisposition, and in particular APOE ε4 homozygosity, has been established as a strong predictor of haemorrhagic risk. [12] Evidence linking vascular risk factors and amyloid burden with ARIA-H risk remains more limited and less consistent in the current literature.

However, these predictors are typically assessed at baseline and used to estimate risk over the duration of a study, often focusing on outcomes at later time points. As a result, such predictors are typically treated as static risk factors, with haemorrhagic risk accordingly conceptualised as a fixed property, rather than a dynamic process with temporal fluctuations. Understanding and modelling the time-varying nature of ARIA-H therefore represents an underexplored opportunity with potential to improve clinical decision-making and risk stratification.

We therefore explored whether a longitudinal, temporally explicit approach to ARIA-H prediction better characterises the dynamics of haemorrhagic risk. Rather than treat ARIA-H as a fixed risk, modelling ARIA-H in discrete intervals between MRI scans allows for analysis of emerging haemorrhagic lesions over time. This approach enables the simultaneous consideration of current accumulation (“how much damage exists”), and the lagged variable of recent accumulation (“how active the process is”), which can be conceptualised as reflecting the current pathological state and trajectory respectively. Distinguishing between these processes may allow for improved risk prediction and provide insight into the underlying processes and mechanisms driving ARIA-H.

In this study, we utilised data from the A4 trial, representing a large and well-monitored cohort of cognitively unimpaired participants with elevated amyloid. [14] In the A4 study, participants were randomised to treatment with solanezumab or placebo. As solanezumab had limited effects on cerebral amyloid burden, this cohort provides a robust sample for assessing the temporal dynamics of ARIA-H in a setting with relatively limited influence from active amyloid clearance. Using a person-interval dataset constructed from consecutive MRI assessments, we evaluated the temporal evolution and the role of temporally dynamic predictors in the short-term prediction of haemorrhagic risk. Analyses were conducted across time-to-event and all-event approaches to assess variation across these analytic cohorts. We specifically evaluated whether recent microhaemorrhage accumulation independently predicts short-term ARIA-H risk beyond baseline risk factors and current lesion burden.

## 2 Methods

### 2.1 Study design

We conducted a secondary data analysis of the A4 study, which enrolled cognitively unimpaired individuals with elevated amyloid. We conducted a longitudinal analysis of MRI data collected within the study.

Inclusion criteria for the analysis were sufficient data to establish at least one longitudinal value for change in microhaemorrhage count. As the primary analyses included prior-interval microhaemorrhage change as a lagged predictor, the regression models required at least three MRI assessments for inclusion (N=1069 participants). The dataset was segmented into intervals, with the start and end of each interval delineated by MRI scans. MRI scans were interpreted externally by radiologists in the original A4 study.

### 2.2 Person-Interval Dataset Construction

MRI data were structured as a person-interval dataset. For each participant, MRI scans were ordered chronologically, and consecutive scans defined discrete time intervals. Each interval represented the period between two MRI assessments, during which incident haemorrhagic lesions could occur. Participants contributed multiple intervals where applicable. Demographic characteristics of the participants are displayed in Table 1.

**Table 1:** Baseline characteristics of study participants. Data are mean (SD) or n (%). Table includes participants in the person-level analytic cohort used to construct the interval dataset.

| Characteristic | Participants (N=1,140) |
| --- | --- |
| Age, years | 71.9 (4.8) |
| Amyloid burden, Centiloids | 66.0 (32.8) |
| White matter hyperintensity (WMH) | 5.0 (6.3) |
| **Sex, n (%)** |  |
| Female | 679 (59.6%) |
| Male | 461 (40.4%) |
| **Race, n (%)** |  |
| White | 1,074 (94.2%) |
| Black | 27 (2.4%) |
| Asian | 23 (2.0%) |
| Other | 16 (1.4%) |
| **APOE ε4 status, n (%)** |  |
| Non-carrier | 471 (41.3%) |
| Heterozygote | 575 (50.4%) |
| Homozygote | 94 (8.2%) |
| Systolic blood pressure, mmHg | 136.6 (16.1) |
| **Baseline microhaemorrhage count, n (%)** |  |
| 0 | 884 (77.5%) |
| 1 | 181 (15.9%) |
| 2 or more | 75 (6.6%) |
| **Baseline superficial siderosis, n (%)** |  |
| 0 | 1,125 (98.7%) |
| 1 or more | 15 (1.3%) |
| **Treatment assignment, n (%)** |  |
| Solanezumab | 558 (48.9%) |
| Placebo | 582 (51.1%) |

For time-to-event analyses, a dataset was constructed in which participants contributed intervals up to and including the first qualifying event. A separate dataset including all intervals was retained for repeated-interval analyses across the full follow-up period.

### 2.3 Outcome

The study outcome was new ARIA-H in any interval, defined as two or more new microhaemorrhages from the previous interval or any new superficial siderosis from the previous interval. As such, this represents a binary indicator of new haemorrhagic progression between consecutive scans. This threshold was chosen to represent a clinically meaningful increase in haemorrhagic activity.

### 2.4 Exposure and Covariates

We explored both dynamic time-dependent predictors as well as baseline covariates in modelling. Dynamic covariates included change in prior interval microhaemorrhage count, as well as microhaemorrhage count at the start of each interval.

Recent microhaemorrhage accumulation was defined as the change in microhaemorrhage count during the immediately preceding MRI interval. Current microhaemorrhage burden was defined as the microhaemorrhage count at the start of the interval.

Baseline covariates were selected based on biological plausibility and existing literature, and included treatment with solanezumab, APOE ε4 carrier state, age, systolic blood pressure, baseline amyloid, and white matter hyperintensity (WMH) tertile. Systolic blood pressure and amyloid Centiloid were scaled per 10 mmHg and per 10 Centiloids respectively.

### 2.5 Statistical analysis

Discrete-time interval logistic regression was used to model the probability of interval haemorrhagic progression. To account for within-subject correlation arising from repeated intervals from the same participant, cluster-robust standard errors were calculated.

A series of nested models were specified to investigate the relative contributions of dynamic variables to model fit in predicting ARIA-H. Model 1 included only baseline covariates. Model 2 included microhaemorrhage burden at the start of each interval to account for current lesion burden. Model 3 additionally included change in microhaemorrhage burden in the previous interval. Model 4 included baseline covariates and prior-interval MCH change without adjustment for current burden, allowing assessment of the total effect of recent accumulation independent of baseline factors. Model fit was compared using Akaike information criterion (AIC) and Bayesian information criterion (BIC), with lower values indicating better relative fit among models fitted to the same analytic sample. [15,16]

As recent microhaemorrhage accumulation was defined using the immediately preceding interval, dynamic models were restricted to intervals in which this prior-change was available. To account for any time-dependence of ARIA-H, models included MRI interval as a categorical variable. For valid model comparison, nested models were tested on the same analytic sample.

### 2.6 Temporal pattern of ARIA-H incidence

We investigated the temporal dynamics of ARIA-H risk, and whether the risk of developing ARIA-H increases over time across strata defined by subject-level risk factors. The time-to-event dataset was analysed, with 1140 participants prior to the exclusion of missing prior-interval MCH change. We limited the analysis period to include only the first five intervals, according to the MRI scans mandated by the study protocol. [14]

The cohort were stratified into groups based on subject level risk factors including age at baseline, treatment arm, APOE ε4 status, and white matter hyperintensity. We analysed the interval hazard for each stratified group. Cumulative incidence values were derived by estimating cumulative hazard from interval-specific Nelson–Aalen increments. [17,18]

## 3 Results

### 3.1 Study population and dataset construction

Of the 1169 participants with MRI data, 1140 contributed to the interval dataset after excluding participants with insufficient MRI data or missing baseline covariates (Figure 1). In order for the lagged variable of prior interval microhaemorrhage (MCH) change to be defined, a minimum of two intervals with at least three MRI scans were required. The final datasets comprised 1069 participants in the all-event analysis and 1063 participants in the time-to-event analysis.

**Figure 1:**
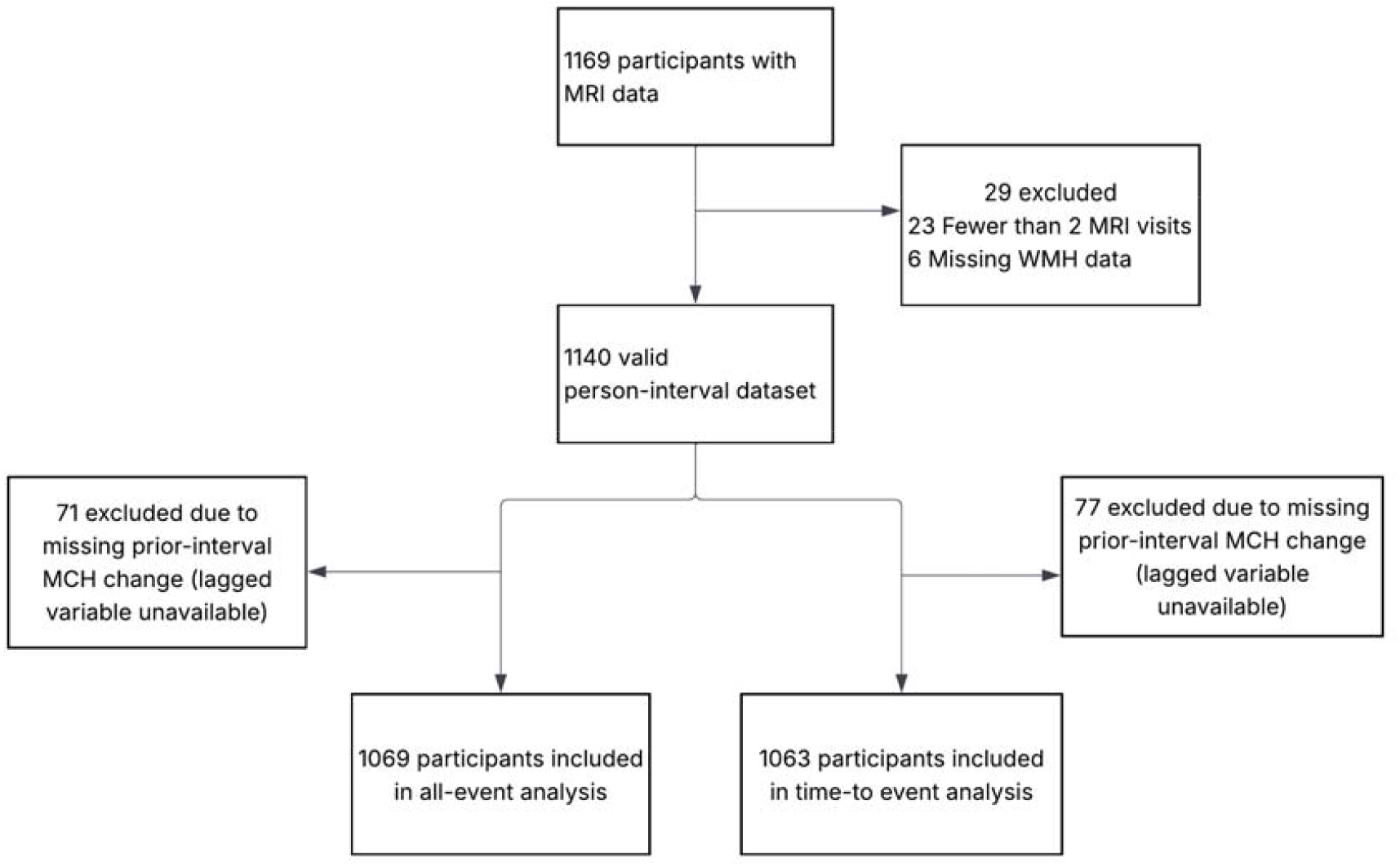
Participants with MRI data were used to construct a person-interval dataset after excluding those with missing data. Two analytic cohorts were defined: an all-interval dataset and a time-to-event dataset. Participants were further excluded due to insufficient interval data to form the lagged prior interval MCH change predictor.

171 ARIA-H events occurred in the all-event analysis from a total of 3,647 intervals and 1,069 participants included. 117 events occurred in the time-to-event analysis from a total of 3,486 intervals and 1,063 participants.

### 3.2 Distribution of ARIA-H events across intervals

Table 2 shows the interval-risk of ARIA-H across successive intervals. Across successive intervals, the number of contributing participants declined from 1,139 in interval 1 to 735 in interval 5. Interval ARIA-H risk increased from 0.4% (95% CI: 0.1-1.0) to 6.8% (95% CI: 5.1-8.9) between interval 1 and interval 5. Correspondingly, person-time incidence rates showed a similar pattern, rising from 1.38 per 100 person-years (95% CI 0.45–3.23) in interval 1 to 8.70 per 100 person-years (95% CI 6.46–11.47) in interval 5. These descriptive findings suggest an increased risk of ARIA-H over time in the cohort studied.

**Table 2:**
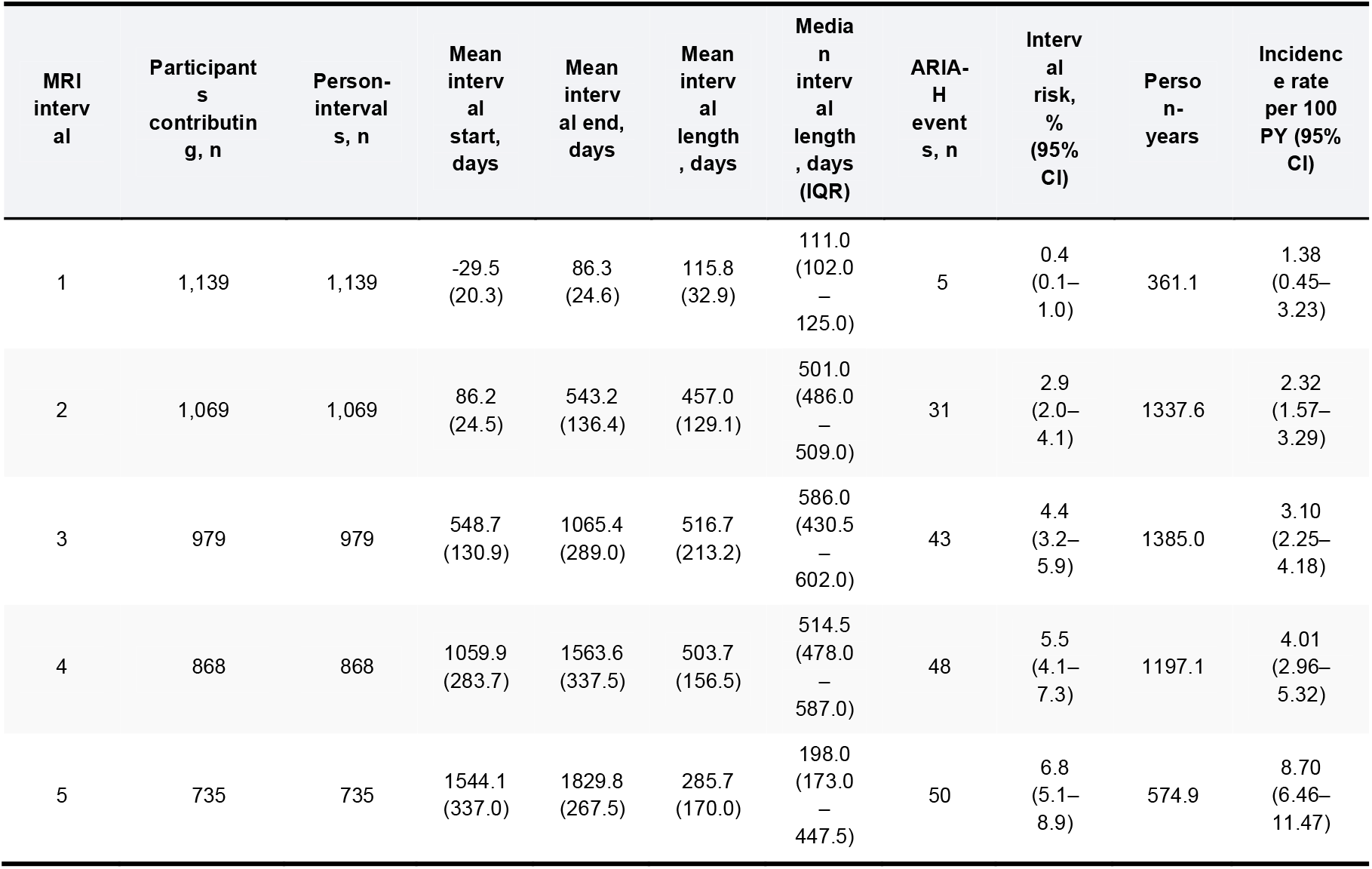
Interval-specific incidence of ARIA-H across MRI intervals. Data are mean (SD), median (IQR), n, or incidence estimates as indicated. Interval risk is the proportion of person-intervals with ARIA-H, with exact binomial 95% confidence intervals. Incidence rates are shown per 100 person-years, with Poisson 95% confidence intervals. CI = confidence interval; IQR = interquartile range; PY = person-years.

Figure 2 shows the cumulative incidence of ARIA-H according to stratified subgroup. Sample sizes varied across analytic subgroups. The clearest separation was observed by APOE ε4 status, with homozygotes demonstrating a higher cumulative incidence throughout the study period. WMH tertile revealed some separation, with low WMH burden associated with a lower ARIA-H risk. Cumulative incidence curves by treatment arm were broadly similar. Age-stratified analysis revealed an increased risk over time in all subgroups, although there was no clear trend towards an increasing risk with age.

**Figure 2:**
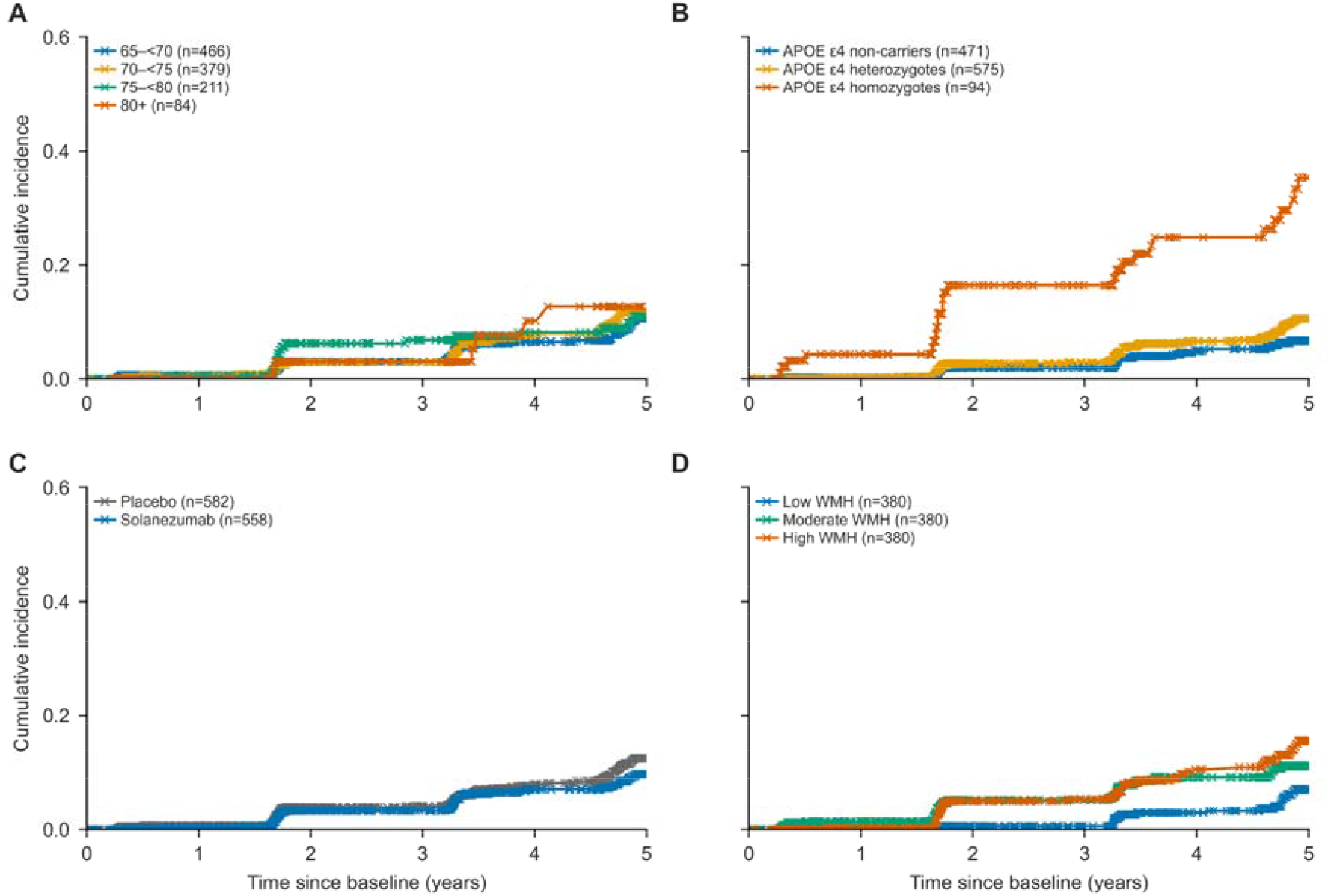
Cumulative incidence of ARIA-H by baseline subgroup. Descriptive cumulative incidence curves are shown according to **(A)** age group, **(B)** APOE ε4 status, **(C)** treatment arm, and **(D)** white matter hyperintensity (WMH) tertile. Cumulative incidence was derived from the Nelson-Aalen estimator.

Descriptive ARIA-H cumulative incidence curves increased across all subgroups over the follow-up period. Although cumulative incidence differed according to subject-level risk factors, particularly APOE ε4 status and WMH burden, upward trajectories were observed across strata, consistent with a time-dependent pattern of ARIA-H risk.

### 3.3 Associations with interval ARIA-H

Figure 3 shows odds ratios for each predictor used in the model, with cluster-robust standard errors to account for repeated intervals for each participant. Both time-to-event associations and all-interval associations were modelled.

**Figure 3:**
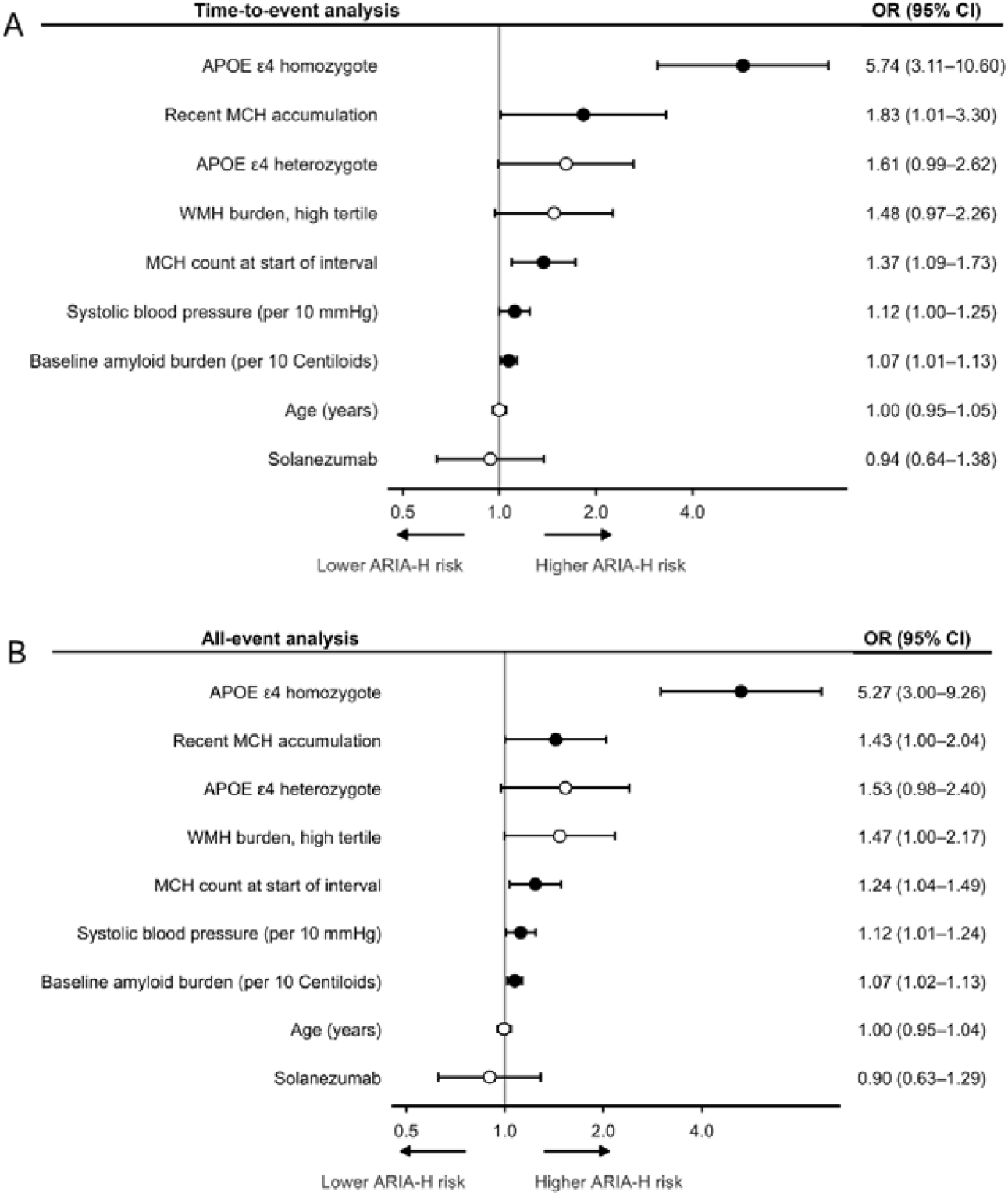
Forest plots show interval ARIA-H odds ratios for time-to-event analysis (A) and all-event analysis (B) Odds ratios (ORs) and 95% confidence intervals (CIs) from clustered discrete-time logistic regression models are shown. Points represent ORs and horizontal lines indicate 95% CIs; filled points denote p<0.05. The x-axis is on a logarithmic scale.

Microhaemorrhage burden at the start of each interval was associated with increased risk of ARIA-H (time-to-event: OR 1.37 95% CI 1.09-1.73; all-event: OR 1.24 95% CI: 1.04-1.49). Recent microhaemorrhage accumulation was associated with increased odds of ARIA-H after adjustment for baseline covariates, current microhaemorrhage burden, and MRI interval (time-to-event: OR 1.83 95% CI 1.01-3.30; all-event: OR 1.43 95% CI: 1.00-2.04).

Consequently, across the two analytic cohorts, both dynamic predictors were associated with a significant increase in the risk of ARIA-H. These findings suggest that both recent haemorrhagic activity and current haemorrhagic burden convey prognostic information beyond baseline risk factors. The associations of both variables with ARIA-H when included in the same model suggest that they capture related but distinct aspects of subsequent haemorrhagic risk.

Across baseline covariates, APOE ε4 homozygosity was associated with the highest odds of ARIA-H in the analysis conducted. Higher baseline systolic blood pressure and amyloid burden were significantly associated with increased haemorrhagic risk in the study period (per 10 mmHg increase in systolic blood pressure: OR 1.12 95% CI 1.00-1.25 and OR 1.12 95% CI 1.01-1.24; per 10 Centiloid increase in amyloid burden: OR 1.07 95% CI 1.01-1.13 and OR 1.07, 95% CI 1.02–1.13 for the time-to-event and all-interval analyses, respectively). Baseline white matter hyperintensity burden showed a positive association that did not reach statistical significance. Odds ratios for treatment with solanezumab, age, and APOE ε4 heterozygosity were non-significant.

Sensitivity analysis was conducted using alternative ARIA-H thresholds. Current microhaemorrhage burden remained consistent with increased risk of ARIA-H. For the broader endpoint of ≥1 new microhaemorrhage or ≥1 new superficial siderosis, current burden was associated with increased risk in both analyses (time-to-event: OR 1.57 95% CI 1.30–1.90; all-event: OR 1.43 95% CI 1.25–1.64). Similar findings were observed for the stricter endpoint of ≥4 new microhaemorrhages or ≥1 new superficial siderosis (time-to-event: OR 1.38 95% CI 1.12–1.70; all-event: OR 1.21 95% CI 1.05– 1.41).

Recent accumulation showed positive but non-significant associations for both alternative endpoints (≥1 endpoint: time-to-event OR 1.35 95% CI 0.89–2.04; all-event OR 1.13, 95% CI 0.88–1.46; ≥4 endpoint: time-to-event OR 1.13 95% CI 0.46–2.80; all-event OR 1.27 95% CI 0.95–1.70). Full results of the sensitivity analysis can be found in Supplementary File 1.

### 3.4 Model performance for prediction of interval ARIA-H

The performance of four discrete-time interval logistic regression models are shown in Table 3. Performance was assessed separately across the time-to-event and all-event analysis. Discrimination was assessed using area under the receiver operating characteristic curve (AUC). Model fit was assessed using Akaike Information Criterion (AIC) and Bayesian Information Criterion (BIC).

**Table 3:** Comparison of discrete-time logistic regression models for prediction of interval ARIA-H Upper panel shows time-to-first-event analysis; Lower panel shows all-event analysis. Models were fitted separately with baseline covariates alone, followed by the introduction of temporal covariates in models 2-4. Model fit was assessed using Akaike Information Criterion (AIC) and Bayesian Information Criterion (BIC), and discrimination using area under the receiver operating characteristic curve (AUC). Abbreviations: AIC, Akaike Information Criterion; BIC, Bayesian Information Criterion; AUC, area under the receiver operating characteristic curve. Confidence intervals for AUC were derived from participant-level bootstrapping.

| Model | Events | AIC | BIC | AUC (95% CI) |
| --- | --- | --- | --- | --- |
| Time-to-event analysis (3,486 intervals; 1,063 participants) |  |  |  |  |
| Model 1: baseline risk only | 117 | 986.6 | 1054.3 | 0.70 (0.69–0.75) |
| Model 2: baseline risk + current burden | 117 | 960.5 | 1034.4 | 0.74 (0.72–0.80) |
| Model 3: baseline risk + current burden + recent accumulation | 117 | 958.0 | 1038.1 | 0.74 (0.71–0.78) |
| Model 4: baseline risk + recent accumulation | 117 | 973.6 | 1047.5 | 0.72 (0.70–0.77) |
| **All-event analysis (3,647 intervals; 1,069 participants)** |  |  |  |  |
| Model 1: baseline risk only | 171 | 1273.7 | 1341.9 | 0.74 (0.72–0.80) |
| Model 2: baseline risk + current burden | 171 | 1153.9 | 1228.3 | 0.80 (0.76–0.85) |
| Model 3: baseline risk + current burden + recent accumulation | 171 | 1145.2 | 1225.8 | 0.80 (0.77–0.84) |
| Model 4: baseline risk + recent accumulation | 171 | 1160.7 | 1235.1 | 0.78 (0.75–0.82) |

Inclusion of current microhaemorrhage burden was associated with a modest increase in model discrimination compared to baseline covariates alone. In the time-to-event analysis, AUC improved from 0.70 in Model 1 to 0.74 in Model 2; in the all-event analysis, AUC improved from 0.74 in Model 1 to 0.80 in Model 2. Corresponding reductions in AIC and BIC indicated improved model fit across both analyses.

Further inclusion of recent microhaemorrhage accumulation in addition to current burden resulted in minimal changes to model discrimination. In the time-to-event analysis, the addition of recent accumulation resulted in a small reduction in AIC (ΔAIC=2.5), but no improvement in BIC. In the all-event analysis, both AIC and BIC improved following incorporation of recent accumulation.

Model 4 included only baseline covariates and recent accumulation to ascertain the independent effect of recent accumulation. Model fit improved and discrimination showed a modest increase in comparison to the baseline risk model.

## 4 Discussion

In this longitudinal analysis of the A4 study, both recent microhaemorrhage accumulation and current microhaemorrhage burden were independently associated with ARIA-H risk. Our results support the conceptualisation of ARIA-H risk as a dynamic property, rather than as a fixed quantity observed prior to trial enrolment or treatment initiation. However, sensitivity analyses provided more robust support for current microhaemorrhage burden as an interval-level predictor, while the association with recent accumulation was less consistent across alternative outcome definitions.

Although both current burden and recent accumulation can be considered as similar entities, our findings suggest that they capture independent properties of haemorrhagic progression, with current burden representing process state and accumulation representing trajectory. Their associations suggest that they capture distinct but complementary dimensions of short-term risk. Importantly, these associations were observed even after accounting for MRI interval, suggesting that recent accumulation was not merely a proxy for later follow-up time.

The results of model fit metrics provide further support for the utility of dynamic variables. Inclusion of current burden produced the clearest improvement in performance, with lower AIC and BIC and modestly higher AUC in both the time-to-event and all-event analyses. This is consistent with the interpretation that current microhaemorrhage burden captures a component of haemorrhagic susceptibility. [9]

Adding recent microhaemorrhage accumulation to a model already containing current burden produced mixed effects on model performance. In the time-to-event analysis, AIC improved, whereas BIC increased and AUC remained unchanged, suggesting limited additional discriminative value or model fit improvement according to the latter more heavily penalised fit criteria. In contrast, for the all-event analysis, both BIC and AIC improved whereas AUC remained unchanged. Taken together, these results suggest a trend towards improved fit with the incorporation of recent accumulation, with stronger support in the all-event analysis than the time-to-event analysis.

Sensitivity analyses using both broader and stricter ARIA-H definitions support the interpretation that there is more robust evidence supporting the association of current microhaemorrhage burden than recent microhaemorrhage accumulation. Current microhaemorrhage burden remained independently associated with ARIA-H across thresholds, whereas recent microhaemorrhage accumulation showed directionally positive but non-significant associations under alternative thresholds.

Sensitivity analyses were not conducted for microhaemorrhage thresholds above ≥4 due to insufficient sample size under such thresholds, and subsequent model instability. The sensitivity analyses should be interpreted cautiously. Using the lower threshold of ≥1 new microhaemorrhage or ≥1 new superficial siderosis in any interval resulted in a significantly higher event count (time-to-event analysis: 403 events, all-event analysis: 482 events), although this presents its own limitations. Using a lower threshold may increase the relative effect of stochastic variables such as variation in image reporting and acquisition. The higher threshold sensitivity analysis of ≥4 new microhaemorrhages or ≥1 new superficial siderosis in any interval is limited by a relatively smaller sample size (time-to-event analysis: 50 events, all-event analysis: 86 events).

The secondary outcomes of static risk factors revealed several results well established in existing literature. Unsurprisingly, APOE ε4 homozygosity was the strongest predictor of ARIA-H among participants in our analysis. The effects of age, APOE ε4 heterozygosity, and solanezumab were non-significant. Higher baseline WMH trended towards an increased risk, although did not reach significance in either analytic cohort. Amyloid burden and baseline systolic blood pressure showed modest positive associations with ARIA-H risk. These findings are biologically plausible given the association of amyloid burden with angiopathy, and the potential for chronic hypertension to increase haemodynamic stress on already fragile amyloid-affected small vessels and promote endothelial and blood–brain barrier dysfunction. [2,4, 19–22]

Biologically, recent microhaemorrhage accumulation may represent a period of heightened vascular instability, rather than chronic lesion accumulation at a constant rate. In the context of cerebral amyloid angiopathy, possible explanations for this include transient worsening of vessel-wall fragility, blood–brain barrier and perivascular dysfunction, or local inflammatory activity, each of which could increase the likelihood of further short-term leakage. [20, 23–25] The observation of this pattern in the A4 study, in which amyloid clearance was limited and solanezumab was not associated with increased haemorrhagic risk, suggests that it may reflect an underlying disease-related process rather than a purely treatment-induced effect. [14] However, it remains unclear whether the observed associations reflect amyloidosis as opposed to microvascular changes related to non-amyloid small-vessel disease. The extent to which amyloid mobilisation amplifies this process remains an important area for future research.

Our results have implications for ARIA-H risk-stratification on a clinical level in the context of anti-amyloid therapies, where clinical decisions depend on assessment of short-term haemorrhagic risk. [8,9] Dynamic measures may have utility in complementing established baseline risk factors for predicting short-term ARIA-H risk by identifying periods of vascular vulnerability. If validated in clinical cohorts receiving effective amyloid-clearing therapies, dynamic risk metrics may guide decision-making surrounding treatment discontinuation or suspension, recommended frequency of MRI monitoring, and individual-level risk counselling. The associations observed for amyloid burden and systolic blood pressure suggest a role for integrating these baseline risk factors in modelling ARIA-H risk, as well as their use in clinical risk-stratification. Although this study did not aim to build a prediction model, and the results do not yet support direct clinical implementation, they suggest that dynamic risk metrics may have a role in future risk stratification and clinical decision making.

### 4.1 Comparison to existing literature

Prior studies have primarily focused on baseline risk factors for ARIA-H. In the existing literature, the primary analytic approaches include observation at the start and endpoint of studies, as well as time-to-event analysis. [11,13] Baseline microhaemorrhage count, although not the primary focus of this study, has been established as a predictor of future haemorrhagic risk. [2,11] To our knowledge, current studies have largely focused on baseline predictors, and we did not identify any studies explicitly modelling current lesion burden and recent microhaemorrhage accumulation as separate time-varying predictors of short-term ARIA-H risk.

Our findings are consistent with the existing literature on genetic risk factors, with APOE ε4, especially homozygosity, being a strong risk factor for ARIA-H. [2,6] The association between amyloid burden and ARIA-H is theoretically plausible, although evidence supporting the association is variable. [3,12,13] Hypertension has been suggested as a potential risk factor for ARIA more generally (including ARIA-E) in the current literature, although evidence linking hypertension with ARIA-H in the context of amyloid-therapeutics is limited. [6,26,27,28]

Although Shirzadi et al. (2025) also analysed ARIA-H in the A4 study, their design differed from the present work. Their primary analysis related baseline factors to haemorrhagic lesion status at the last available MRI, rather than using a discrete time-interval framework to model scan-to-scan risk. Consistent with the present results, APOE ε4 homozygosity emerged as a strong risk factor. However, whereas Shirzadi et al. reported an independent association between elevated white matter hyperintensity burden and haemorrhagic lesion emergence, WMH in the present analysis showed only a positive but non-significant association with interval ARIA-H risk. These differences may reflect the different analytic approaches, modelling implementation, or inclusion criteria. [13]

### 4.2 Strengths and limitations

Strengths of this study include the robust dataset; the A4 study allows for analysis of a well-monitored cohort of participants according to a pre-defined protocol. [14] Furthermore, the study implemented a standardised protocol for central reading of MRI scans, improving the validity of analyses conducted.

Our longitudinal discrete time-interval approach to dataset construction and analysis provides a coherent analytic framework in which ARIA-H can be conceptualised as an event between two time points, rather than assuming a continuous time variable. We conducted both time-to-event and all-event analyses, allowing for validation across two distinct cohorts. Our uncertainty estimates utilised clustered uncertainty estimation to account for repeated participant intervals in the dataset.

However, several limitations should be acknowledged. It should be noted that the A4 study was conducted in a selected, predominantly white, pre-symptomatic sample of participants with preclinical Alzheimer’s disease. [14] As such, our findings may not fully generalise to symptomatic individuals or routine clinical cohorts.

The potential effect of amyloid-clearing therapeutics on our results remains unclear. Further research is needed to demonstrate whether our results translate to cohorts treated with contemporary anti-amyloid therapies. Our outcome definition was chosen as a clinical benchmark for significant ARIA-H, although other endpoint thresholds could be justified, likely with differing results. We note that ARIA-H is measured between MRI scans, and differences in interval spacing may introduce uncertainty in our results. Even with central MRI reading, some variability exists in image reporting, with the additional variable of MRI magnetic field strength potentially affecting haemorrhagic lesion readings. [29,30] While independent associations and improvements in model fit were observed, improvements in model discrimination were modest throughout the analysis. Although the development of a validated prediction model is beyond the scope of this study, we acknowledge that external validation would provide evidence for the generalisability of our results. Finally, as a secondary observational analysis, this study cannot establish causal relationships between covariates and subsequent ARIA-H.

### 4.3 Conclusions

Our results suggest that both recent microhaemorrhage accumulation and current microhaemorrhage burden are independently associated with ARIA-H in the primary endpoint analysis. These findings support the conceptualisation of ARIA-H as a dynamic process in which current lesion burden, alongside recent lesion activity, may improve the clinical prediction of haemorrhagic risk. Given the recent introduction of amyloid-clearing therapies into clinical practice, both improving the understanding of haemorrhagic risk in treated populations, and individual-level temporal prediction of haemorrhagic risk may inform clinical decision-making and monitoring strategies. If validated in suitable cohorts receiving anti-amyloid therapies, our findings may inform a more individualised and dynamic approach to ARIA-H risk assessment and monitoring.

## Clinical trial registration

Clinical trial registration: Clinical Trial of Solanezumab for Older Individuals Who May be at Risk for Memory Loss (A4) (NCT02008357)

## Data availability statement

The data that support the findings of this study are available to researchers following approval through the Alzheimer’s Clinical Trial Consortium A4/LEARN data portal: https://www.a4studydata.org

## Ethics statement

This study was a secondary analysis conducted in accordance with the applicable data-use agreement. Ethical approval was granted by the Faculty of Medicine Ethics Committee (ERGO:112166).

## Acknowledgements

The A4 Study was a secondary prevention trial in preclinical Alzheimer’s disease, aiming to slow cognitive decline associated with brain amyloid accumulation in clinically normal older individuals. The A4 Study was funded by a public-private-philanthropic partnership, including funding from the National Institutes of Health-National Institute on Aging, Eli Lilly and Company, Alzheimer’s Association, Accelerating Medicines Partnership, GHR Foundation, an anonymous foundation, and additional private donors, with in-kind support from Avid Radiopharmaceuticals, Cogstate, Albert Einstein College of Medicine and the Foundation for Neurologic Diseases. The companion observational Longitudinal Evaluation of Amyloid Risk and Neurodegeneration (LEARN) Study was funded by the Alzheimer’s Association and GHR Foundation. The A4 and LEARN Studies were led by Dr. Reisa Sperling at Brigham and Women’s Hospital, Harvard Medical School, and Dr. Paul Aisen at the Alzheimer’s Therapeutic Research Institute (ATRI) at the University of Southern California. The A4 and LEARN Studies were coordinated by ATRI at the University of Southern California, and the data are made available under the auspices of Alzheimer’s Clinical Trial Consortium through the Global Research & Imaging Platform (GRIP). The complete A4 Study Team list is available at: https://www.actcinfo.org/a4-study-team-lists/. We would like to acknowledge the dedication of the study participants and their study partners who made the A4 and LEARN Studies possible.

## Supporting information

Supplementary File 1, Sensitivity Analysis

